# Are HIV treatment clients offered a choice of differentiated service delivery models? Evidence from Malawi, South Africa, and Zambia

**DOI:** 10.1101/2024.12.09.24317166

**Authors:** Idah Mokhele, Vinolia Ntjikelane, Nancy Scott, Jeanette L. Kaiser, Allison Morgan, Amy Huber, Oratile Mokgethi, Timothy Tchereni, Wyness Phiri, Aniset Kamanga, Prudence Haimbe, Priscilla Lumano-Mulenga, Rose Nyirenda, Sophie Pascoe, Sydney Rosen

## Abstract

**Purpose:** Differentiated service delivery (DSD) models for antiretroviral therapy (ART) for HIV are intended to increase patient-centeredness, a concept that incorporates patient choice of service delivery options. We explored choice in DSD model enrollment at 42 public sector clinics in Malawi, South Africa, and Zambia.

**Methods:** From 09/2022-05/2023, we surveyed ART clients and healthcare providers, asking ART clients if they had a choice about DSD model enrollment and providers about their practices in offering choice. We estimated risk differences for ART clients’ self-reported offer of choice using a Poisson distribution with an identity link function and report risk differences. We thematically analyzed open-ended questions and report key themes.

**Results:** We enrolled 1049 clients (Malawi 409, South Africa 362, Zambia 278) and 404 providers (Malawi 110, South Africa 175, Zambia 119). The proportion of clients indicating that they had been offered a choice ranged from 4% in Malawi to 17% in Zambia to 47% in South Africa. Few clients (Malawi 10%, South Africa 19%, Zambia 13%) reported they had actively asked to enroll in a DSD model, but a majority (Malawi 66%, South Africa 80%, Zambia 59%) indicated they consented to model enrollment, even if enrollment was not presented as a choice. Over 90% of clients in all three countries reported that they were happy to be enrolled in their current DSD model. Among providers, 64% in Malawi, 80% in South Africa, and 59% in Zambia said they offered clients the choice to enroll in DSD or remain in conventional care.

**Conclusions:** As of 2023, relatively few ART clients in Malawi, South Africa, and Zambia said they were offered a choice about enrolling in a differentiated service delivery model, despite most providers reporting offering a choice. The value of patient choice in improving clinical outcomes and satisfaction should be explored further.

**Plain-Language Summary:** *Purpose:* Differentiated service delivery (DSD) models for antiretroviral therapy (ART) enhance patient-centeredness by offering choices in service delivery options. We explored choice in DSD model enrollment at 42 public sector clinics in Malawi, South Africa, and Zambia.

*Methods:* From 09/2022-05/2023, we surveyed ART clients and healthcare providers, asking clients whether they had a choice about DSD model enrollment and providers whether they offered this choice to their clients. We analyzed the data to identify differences in how clients reported being offered a choice.

*Results:* 1,049 clients participated: 409 from Malawi, 362 from South Africa, and 278 from Zambia, alongside 404 providers (110 in Malawi, 175 in South Africa, and 119 in Zambia). 4% of clients in Malawi, 17% in Zambia, and 47% in South Africa reported being offered a choice to enroll in a DSD model. Few actively sought to join a DSD mode (Malawi 10%, South Africa 19%, and Zambia 13%)—but many consented to enroll even when not explicitly offered (Malawi 66%, South Africa 80%, Zambia 59%). Over 90% of all clients were happy with their current DSD model. Among providers, 64% in Malawi, 80% in South Africa, and 59% in Zambia offered clients the choice to enrol in a DSD model or remain in conventional care.

*Conclusions:* Most ART clients in our study did not report being offered a choice to join a DSD model despite providers claiming they offered one. Further research is needed to understand how offering choices could improve health outcomes and patient satisfaction with care.

## Introduction

Differentiated service delivery (DSD) models for HIV treatment aim to increase the extent to which service delivery is client-centered, a term broadly defined as care that is holistic and responsive to individual needs. ^1–3^ An important element is shared decision-making between providers and clients that empowers clients and can improve outcomes. ^4^ DSD adjusts service delivery to meet the needs of treatment clients in terms of the location, frequency, and other characteristics of interactions with the healthcare system. ^5^ For DSD for HIV treatment, healthcare systems are expected to offer treatment clients information about different options and a choice of available service delivery models so that individual clients can select the model that best meets their needs.^6^

Many sub-Saharan African countries have actively implemented differentiated service delivery for HIV treatment since 2016, when World Health Organization guidelines first recommended this approach. ^3^ The years since then have seen countries experiment with, adopt, scale up, and retire various models of service delivery as they have gained experience with differentiation and the benefits and costs of specific DSD models. While DSD implementation continues to evolve, many countries are gradually converging on the widespread use of a few models for “established” or stable ART clients who have been on treatment for at least 6 months and are virally suppressed. These include multi-month dispensing of ART that reduces the frequency of clinic visits; facility-based fast-track models that allow clients to refill prescriptions without waiting in regular clinic queues; community-based medication pickup points; and, to a lesser extent, group or club models and home medication delivery. In most countries, clients may also opt to remain in conventional (undifferentiated) care, which generally requires ^4–6^ full clinic visits and medication refills per year.

Each of these models offers established ART clients a different set of service delivery characteristics, benefits, and costs and should thus be preferred by different groups of clients, depending on their own circumstances and constraints.^7^ While national guidelines in many countries specify a range of DSD models to be made available ^8–11^, the extent to which clients are offered a choice of models, or of any differentiated model versus remaining in conventional care, is unclear. As part of a survey of the benefits and costs of DSD models for ART, we asked ART clients enrolled in DSD models in Malawi, South Africa, and Zambia whether they had been offered an opportunity to choose their model of care and explored which characteristics of clients and facilities were associated with being offered a choice. In addition, we asked providers whether they offer clients a choice between models and, if not, their reasons for not providing this choice.

## Materials and methods

### Study sites

The AMBIT Project’s SENTINEL 2.0 survey was the second round of a repeated, cross-sectional, interviewer-administered survey delivered to a sample of ART clients and healthcare workers at 12 public sector clinics in Malawi, 18 in South Africa, and 12 in Zambia during the period from September 2022 to May 2023. The study design has previously been published ^12^, and study sites are briefly described in Supplementary Table 1.

The study sites reported offering varying combinations of models of care, depending on national guidelines, facility size and resources, and, in the case of Malawi and Zambia, the presence of a nongovernmental partner organization that contributed equipment, staff, and other resources for specific models of care. For clients meeting criteria to be considered established on ART, medication dispensing intervals ranged from 2-6 months per pickup. Facility-based, six-month dispensing of medications was rapidly expanding in Malawi and Zambia at the time of the survey, and facilities in each country also offered community medication pickup points and various population-specific models such as teen/youth clubs and mother-infant pair clinics. South Africa offers three “less intensive” models for ART clients established on treatment, namely facility-based medication pickup points, external (out-of-facility) medication pickup points, and adherence clubs. (In South Africa, these options are known as Differentiated Models of Care (DMOC) and, as less intensive models, classified under Repeat Prescription Collection strategies (RPCs).) Although adherence club models and patient-led group models of care were originally popular, at the time of study enrolment, club and group models had been scaled back significantly because of COVID-19, and we did not encounter these models at most of the study sites. Supplementary Table 1 describes the lists of commonly offered models of care found in each country when the survey was administered.

For the current analysis the models of care were combined into three categories, based on a commonly used taxonomy of DSD models.^5^

- Facility-based individual models: These included facility medication pickup points, fast-track services, 6-month multi-month dispensing (MMD), and high-intensity models for ART clients who were not yet established on ART.
- Facility-based group models: This category comprised adherence clubs and family models, which were structured to support group-based care within the facility setting.
- Community-based group or individual models: These were either individual and group-based models implemented in the community. They encompassed community-based pickup points and home-delivery options for individuals. The group-based models, implemented within community settings, included patient or provider-led adherence or outreach groups.

### Study populations and enrollment

We enrolled two discrete populations into SENTINEL 2.0. The first study population comprised adult ART clients with 6 or more months’ experience on ART, at which point all participants had sufficient time on ART to be eligible for DSD enrollment, if they met other criteria for being designated as "established." At each study site, up to 10 clients per active model of care were enrolled. (Although clients remaining in conventional care (not enrolled in any DSD model) were also recruited for SENTINEL 2.0, they were not asked questions about DSD model choice and were not included in the analysis reported here. Clients were enrolled in the survey sequentially as they arrived at the facilities for routine HIV-related care. Following written informed consent, they were administered a questionnaire by a study research assistant.

The second study population consisted of up to 10 healthcare providers at each facility. This group included three main categories:

- Healthcare Professionals: This includes nurses, doctors, and clinicians who are involved in patient care and clinical decision-making.
- Lay Health Workers: This category includes lay counsellors, peer educators/ navigators, and community health workers who provide essential support and education to patients.
- Support and Administrative Staff: This group includes data clerks, data capturers, and other administrative personnel who support the operational aspects of healthcare delivery.

Eligible providers had been working at the facility for a minimum of six months and self-reported being involved in implementing DSD models. Potential respondents were referred to the study team by the facility manager and were asked for written informed consent. They were then interviewed by a research assistant using a structured questionnaire. Further details on enrollment procedures for both study populations have previously been published. ^12^

### Survey questions regarding choice of DSD model

Survey instruments for both populations were designed by the study team and included quantitative and open-ended questions aimed at understanding clients’ experiences and providers’ perspectives. Clients were asked if they were offered a choice of model enrollment; if they had asked to be enrolled in their current model; if they were happy to be enrolled in their current model; and whether they would prefer a different model. Providers were asked two questions concerning the offer of choice to clients. The first question inquired whether providers offered established clients the option to enrol in a DSD model or not; if they chose not, they would remain in conventional (non-DSD) care. The second question asked whether they provided established clients with a choice among the available DSD models at the facility and, if not, their reasons for not offering this choice.

The survey also collected information on factors that could affect the offer of choice of DSD model enrollment among ART clients, including patient-related factors such as age and sex and duration of time the client had been on ART medication. Facility characteristics such as location (urban or rural) and size were also recorded. Facility size was based on the total number of patients on ART during the study enrolment period and was classified into three categories: facilities with 1,000-2,000 clients remaining on ART, facilities with 2,000-4,000 clients, 4,000-6,000 and facilities with more than 6,000 clients. The exact language of the questions used in each survey is included in Supplementary files 1 and 2.

### Data analysis

We first describe participant characteristics for each population using proportions, frequencies, means with standard deviations, and medians with interquartile ranges (IQR), as appropriate. For the client survey, we report the proportions of respondents indicating that they were or were not offered a choice and the characteristics of clients, healthcare facilities, and DSD models that are associated with being offered a choice. We estimated the risk differences (RD) for self-reported offer of DSD model choice among study participants using a Poisson distribution with an identity link function. We adjusted for duration on ART, country, age, sex, facility size, and clinic locality and report adjusted risk differences (ARD) and 95% confidence intervals (CI).

For the provider survey, we describe provider characteristics using proportions, frequencies, means with standard deviations, and medians with interquartile ranges (IQR), as appropriate. We report the proportion of providers who reported offering ART clients a choice in DSD model participation. Open-ended questions from the provider survey were used to develop the codebook *a priorie.* The codebook was refined after a reading of responses. All questions were then coded using Excel and analyzed thematically. Results were compared across countries and by respondent type, then triangulated with quantitative findings. Emerging themes were summarized and are presented with illustrative quotes which were lightly edited for clarity when needed.

### Ethics

The study protocol was reviewed and approved by the Human Research Ethics Committee of the University of Witwatersrand in South Africa (protocol M210241), the Boston University Institutional Review Board (IRB) (protocol H-41402), the National Health Science Research Committee (NHSRC) in Malawi (protocol 21/03/2672), and ERES Converge Institutional Review Board in Zambia (protocol 2021-Mar-012). All participants provided written informed consent.

## Results

### ART client study population

A total of 409, 362, and 278 ART clients were enrolled in Malawi, South Africa, and Zambia, respectively. Demographic and socioeconomic characteristics of participants and the DSD models in which they were enrolled are presented in Table 1. Consistent with overall ART uptake in sub-Saharan Africa, roughly two-thirds of participants were female. Unemployment was high in South Africa; informal employment was the most common occupation in Malawi and Zambia. Most participants had already been on ART for more than 5 years at the time of the survey.

**Table 1.** Characteristics of ART client study population (n=1049)

| Characteristic | Malawi | South Africa | Zambia | Total |
| --- | --- | --- | --- | --- |
| N (row percentage)* | 409 (39) | 362 (35) | 278 (26) | 1049 (100) |
| Age (median, IQR) | 33 (22-43) | 41 (35-48) | 41 (22-51) | 38 (28-47) |
| Sex (% female) | 296 (72) | 284 (78) | 181 (65) | 761 (73) |
| Highest level of education completed |  |  |  |  |
| <Grade 12 | 258 (63) | 180 (50) | 153 (55) | 591 (56) |
| ≥Grade 12 | 151 (37) | 182 (50) | 125 (45) | 458 (44) |
| Employment status |  |  |  |  |
| Formal employment | 13 (3) | 101 (28) | 29 (10) | 143 (14) |
| Informal employment | 227 (56) | 103 (28) | 144 (52) | 474 (45) |
| Unemployed | 80 (20) | 149 (41) | 53 (19) | 282 (27) |
| Student/trainee | 89 (22) | 9 (2) | 52 (19) | 150 (14) |
| Number of years on ART (self-report) |  |  |  |  |
| Median (IQR) | 8 (4-13) | 7 (5-11) | 9 (6-14) | 8 (5-12) |
| 1-5 years | 126 (31) | 85 (23) | 50 (18) | 261 (25) |
| 5-10 years | 107 (26) | 162 (45) | 98 (35) | 367 (35) |
| ≥10 years | 176 (43) | 115 (32) | 130 (47) | 421 (40) |
| DSD model enrollment |  |  |  |  |
| Facility-based individual | 298 (73) | 145 (40) | 133 (48) | 576 (55) |
| Facility-based group | 91 (22) | 11 (3) | 88 (32) | 190 (18) |
| Community-based group or individual | 20 (5) | 206 (57) | 57 (21) | 283 (27) |

\*The SENTINEL study also surveyed ART clients who were not enrolled in DSD models but remained in conventional care, but these ART clients were not asked about choices of service delivery models. Here, we report only on those enrolled in DSD models.

### ART clients’ self-report of offers of choice in model participation

Figure 1 illustrates the share of participants in each country who indicated that they had asked to be enrolled in their current model of care, provided consent for enrollment (written or verbal), were given a choice about participating in their current model, and were happy to be enrolled in it. The proportion of participants indicating that they had been offered a choice ranged from 4% in Malawi to 17% in Zambia to 47% in South Africa. Fewer than 14% in each country (Malawi 10%, South Africa 19%, Zambia 13%) indicated they had actively asked to enroll in a DSD model.

**Figure 1a.**
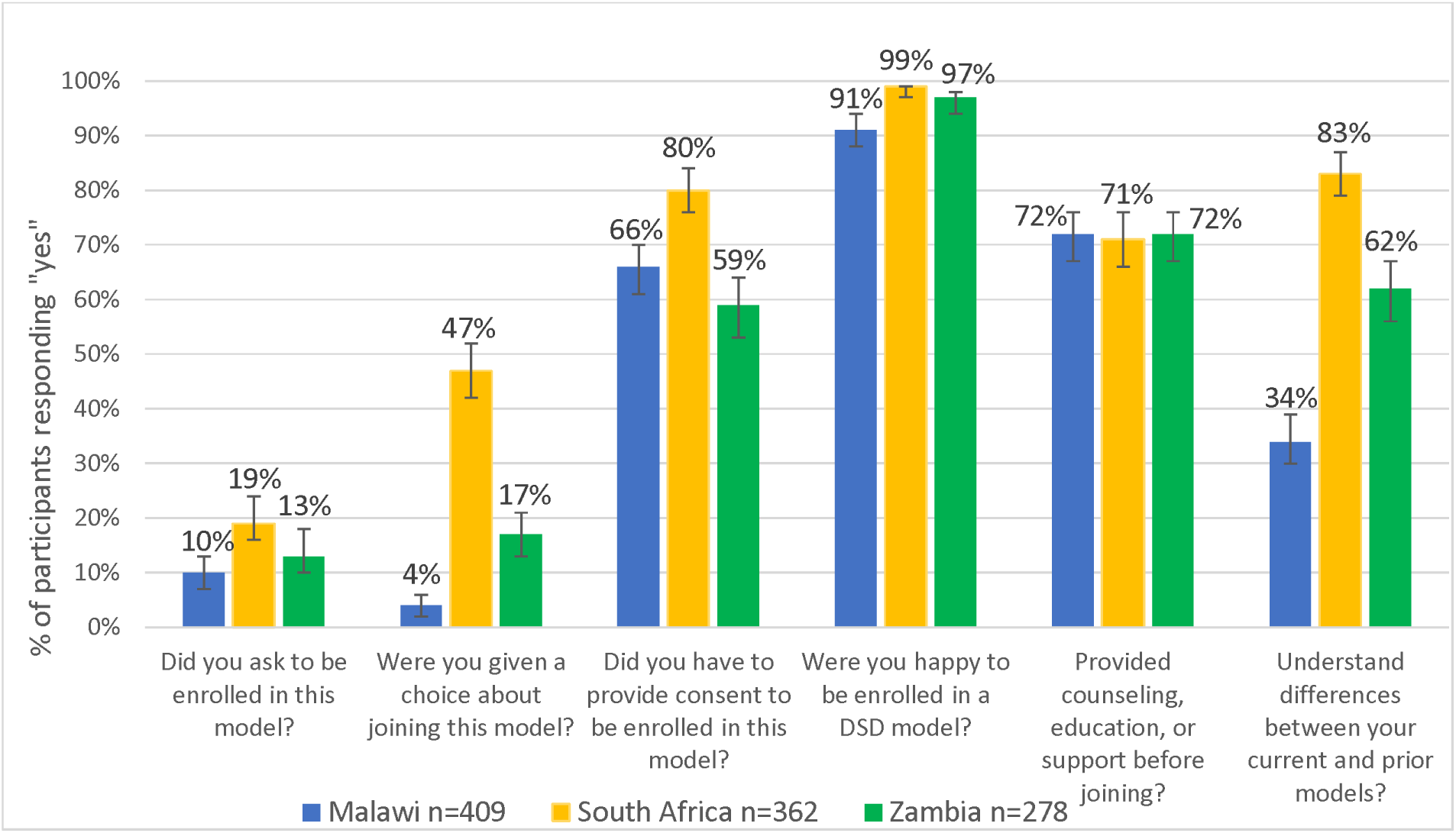
Survey participants’ self-reported offer of choice in enrolling in their current DSD models, by country (n=1,049).

Despite participants’ self-reported lack of choice between DSD models, over two-thirds (Malawi 66%, South Africa 80%, Zambia 59%) indicated they consented to DSD model enrollment (in any model) rather than remaining in conventional care. Among the participants who reported providing consent (723/1049), 22% gave written consent, and 78% gave verbal consent. In Malawi, South Africa, and Zambia, only 1%, 38%, and 4% provided written consent, respectively.

In all three countries, around 70% of individuals reported receiving counselling, education, or support before enrolling in their current model, and 60% said they understood the differences between their current and prior models of care. Overall, 38% discussed their choice of DSD model with their provider at their initial enrollment, and a quarter did so during their most recent clinical visit or drug pickup, though again with variation by country (Figure 1b). Despite the relative lack of choice offered, large majorities, in excess of 90% in all three countries, reported that they were happy to be enrolled in their current DSD model.

**Figure 1b.**
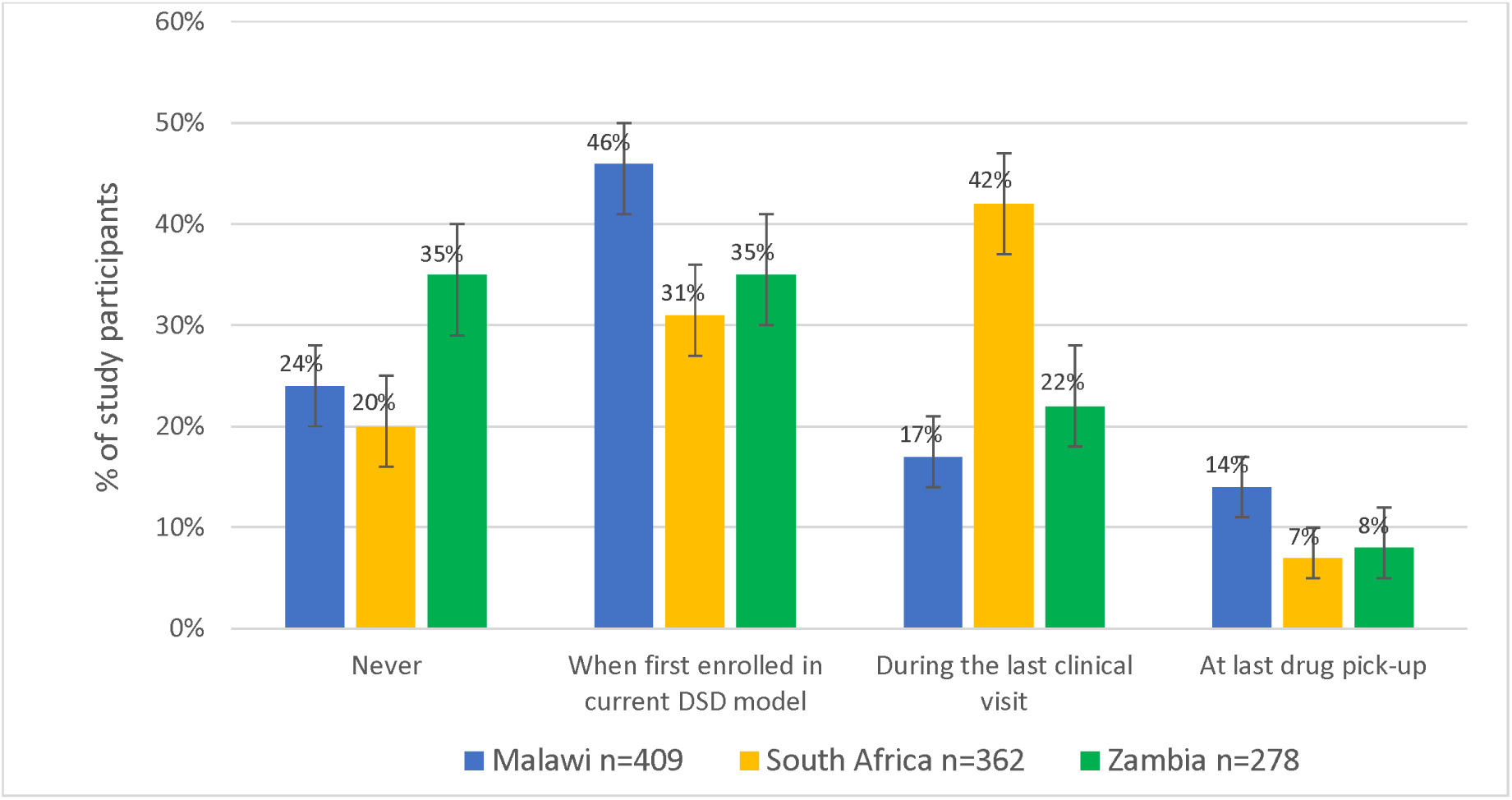
Survey participants self-reported timing of last discussion regarding DSD model choice with provider, by country (n=1,049).

When asked if they were aware of other DSD models currently implemented in their facility and if they preferred any other model to their current model, all participants from South Africa and Zambia and 82% of those from Malawi said they were aware of other models (Table 2). Among those aware of other DSD models, a third said they would have preferred to be in a different model (48% in Malawi, 38% in South Africa, and 19% in Zambia).

**Table 2.** Awareness of and Preferred DSD Model among ART client study population (n=1049)

|  | Malawi<br>(N = 409) | South Africa<br>(N = 362) | Zambia<br>(N = 278) | Total<br>(N = 1049) |
| --- | --- | --- | --- | --- |
| Not aware of other DSD models | 72 (18) | - | - | 72 (6) |
| Aware of other DSD models | 337 (82) | 362 (100) | 278 (100) | 997 (93) |
| **Preferred another DSD model** | **163 (48)** | **136 (38)** | **53 (19)** | **352 (35)** |
| Currently in facility-based individual model | 157 (96) | 51 (38) | 40 (75) | 248 (70) |
| Currently in facility-based group model | 2 (1) | - | - | 2 (1) |
| Currently in community-based group or individual model | 4 (2) | 85 (62) | 13 (25) | 102 (29) |

Qualitatively, among the few patients in Zambia (n=46) and Malawi (n=15) who responded to the open-ended question about having been given a choice, they primarily described it being a single option (i.e. they were offered one of the models) by the provider, with the choice to opt into that model or remain in conventional care. They primarily reported selecting the option that was more convenient, closer to home, or would save time and transport money. Illustrative quotes are included below:

> “[I] was asked if I would prefer the healthcare providers to deliver medication at my home or not.” – Zambia, female patient, age 50-64
>
> “They [the providers] asked for those that were willing to join the community adherence groups.” – Zambia, male patient, age 35-49
>
> “It was the choice to either remain in standard of care or switch to 6MMD.” – Zambia, female patient, age 35-49
>
> “[I was given the choice] to either join adolescent group or not but was told of the benefits that come with joining it.” – Zambia, female patient, age 20-24
>
> “To choose if I can be in 6MMD or not.” – Malawi, male patient, age 25-34
>
> “[I was given the choice] whether to join teen club or to be in standard of care.” – Malawi, male patient, age 15-19
>
> “Choice of whether to accept getting medications from outreach or at the facility.” – Malawi, female patient, age 50-64

Conversely, in South Africa (n=171), patients qualitatively described a much more varied experience. Many stated that providers explained the DSD model options to them as well as the benefits of different models and gave them an option for choosing either to remain on conventional care or opt for a facility or external medication pickup point. Most patients described choosing specifically where they would pick up their medication to make it most convenient for them. Patients also widely discussed choosing their DSD model because it would save them time due to shorter waits and queues and quicker service, which made their experience more convenient and easier. They connected the convenience with the locations of the pickup points (pharmacies, grocery stores, or the clinic), liking that the pickup points were close to either their homes or work, where they could easily walk to and did not require them to miss work. Multiple noted the convenience of someone else being able to pick up medications for them if needed. Illustrative quotes are included below:

> “They [the providers] gave me more options to choose from as to where to collect my medication in a convenient place for me.” – South Africa, female patient, age 35-49
>
> “They [the providers] informed me and asked me nicely if it’s okay with me if they send me to collect medication at Clicks.* They also mentioned the benefits of collecting medication at the External pickup point.” – South Africa, female patient, age 35-49 (*A commercial pharmacy chain in South Africa.)
>
> “I was given different external pickup points within the community, and I choose the one where I’m taking medication now.” – South Africa, male patient, age 35-49
>
> “They [the providers] asked if I prefer to collect at clinic or go to the external pickup point so they did educate me also I was given a choice to choose my preferred method for ART collection they never forced or decided on my behalf.” – South Africa, female patient, age 35-49
>
> “They [the providers] explained the available DSD models which they offer at this facility and that I can choose the one which will be convenient for me.” – South Africa, female patient, age 25-34

### Associations between being offered a choice and client, facility, and model characteristics

Neither age nor sex was associated with the probability of being offered a choice of models of care (Table 3). As mentioned above, choice was much more common in South Africa than in either Zambia or Malawi. Participants enrolled in urban facilities (ARD 0.94 [95% CI: 0.90-0.99]) and in sites with 2000-4000 ART clients were slightly less likely to be offered DSD model enrollment (2000-4000 ART clients vs <2000 ART clients, 0.91 [0.84-0.98]). Clients at high-volume sites, in contrast, were more likely to have requested DSD enrollment (4000-6000 ART clients vs 1000-2000 ART clients, 1.08 [1.00-1.16]), (>6000 ART clients vs 1000-2000 ART clients, 1.12, [1.04-1.22]).

**Table 3.**
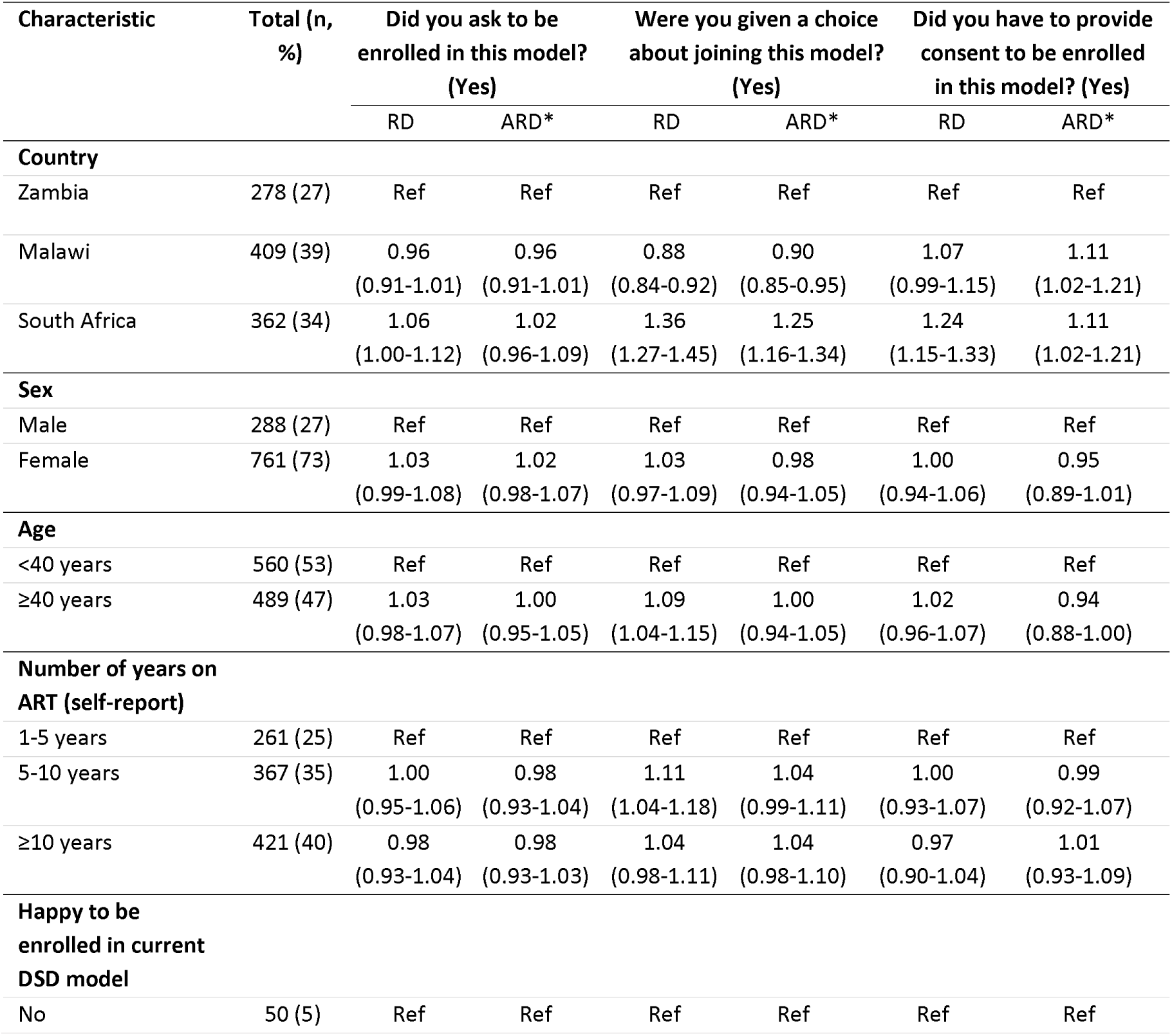
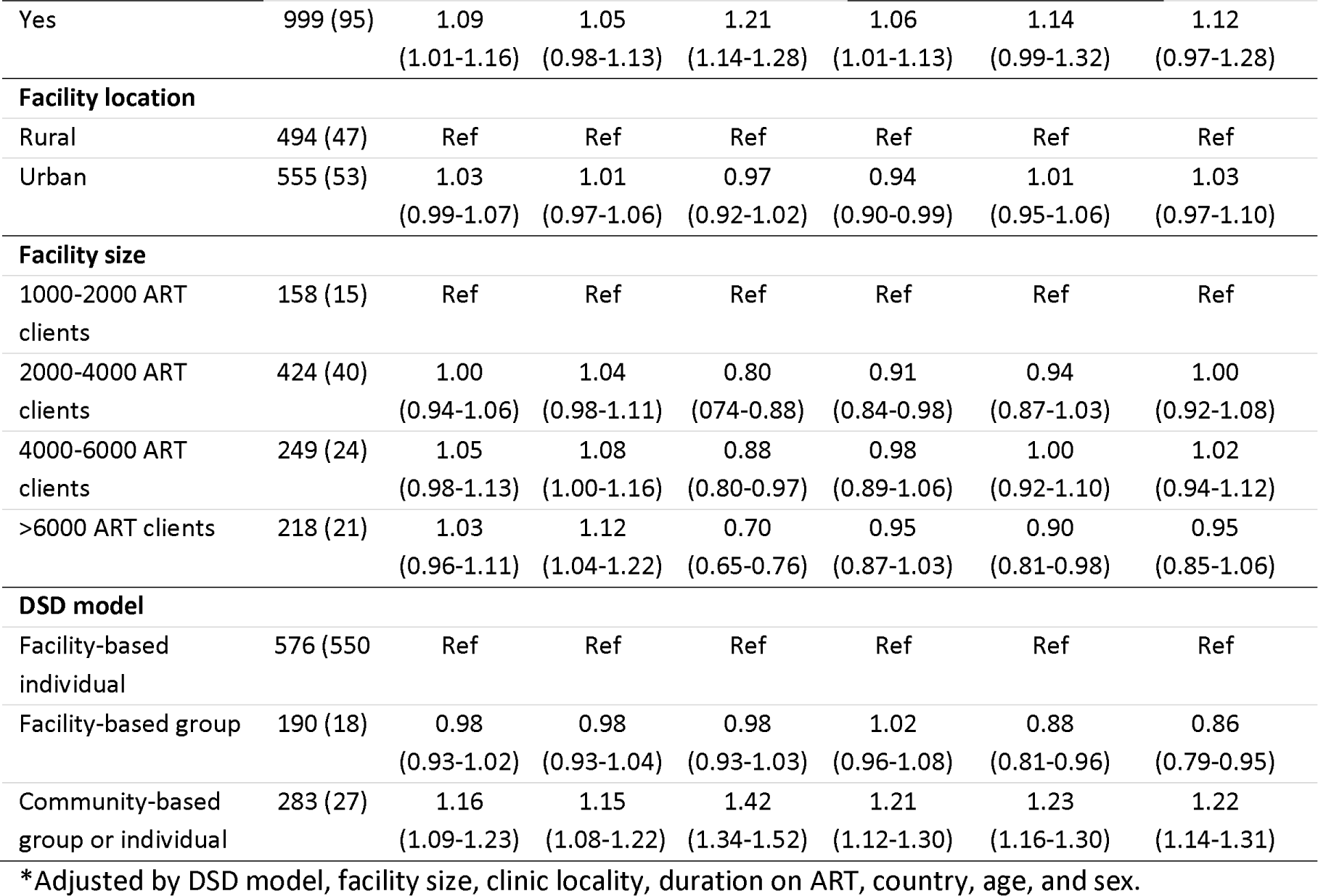
Crude and adjusted risk differences for ART clients’ experiences of DSD model choice (n=1049)

| Characteristic | Total (n, %) | Did you ask to be enrolled in this model? (Yes) |  | Were you given a choice about joining this model? (Yes) |  | Did you have to provide consent to be enrolled in this model? (Yes) |  |
| --- | --- | --- | --- | --- | --- | --- | --- |
|  |  | RD | ARD* | RD | ARD* | RD | ARD* |
| Country |  |  |  |  |  |  |  |
| Zambia | 278 (27) | Ref | Ref | Ref | Ref | Ref | Ref |
| Malawi | 409 (39) | 0.96<br>(0.91-1.01) | 0.96<br>(0.91-1.01) | 0.88<br>(0.84-0.92) | 0.90<br>(0.85-0.95) | 1.07<br>(0.99-1.15) | 1.11<br>(1.02-1.21) |
| South Africa | 362 (34) | 1.06<br>(1.00-1.12) | 1.02<br>(0.96-1.09) | 1.36<br>(1.27-1.45) | 1.25<br>(1.16-1.34) | 1.24<br>(1.15-1.33) | 1.11<br>(1.02-1.21) |
| Sex |  |  |  |  |  |  |  |
| Male | 288 (27) | Ref | Ref | Ref | Ref | Ref | Ref |
| Female | 761 (73) | 1.03<br>(0.99-1.08) | 1.02<br>(0.98-1.07) | 1.03<br>(0.97-1.09) | 0.98<br>(0.94-1.05) | 1.00<br>(0.94-1.06) | 0.95<br>(0.89-1.01) |
| Age |  |  |  |  |  |  |  |
| <40 years | 560 (53) | Ref | Ref | Ref | Ref | Ref | Ref |
| ≥40 years | 489 (47) | 1.03<br>(0.98-1.07) | 1.00<br>(0.95-1.05) | 1.09<br>(1.04-1.15) | 1.00<br>(0.94-1.05) | 1.02<br>(0.96-1.07) | 0.94<br>(0.88-1.00) |
| Number of years on ART (self-report) |  |  |  |  |  |  |  |
| 1-5 years | 261 (25) | Ref | Ref | Ref | Ref | Ref | Ref |
| 5-10 years | 367 (35) | 1.00<br>(0.95-1.06) | 0.98<br>(0.93-1.04) | 1.11<br>(1.04-1.18) | 1.04<br>(0.99-1.11) | 1.00<br>(0.93-1.07) | 0.99<br>(0.92-1.07) |
| ≥10 years | 421 (40) | 0.98<br>(0.93-1.04) | 0.98<br>(0.93-1.03) | 1.04<br>(0.98-1.11) | 1.04<br>(0.98-1.10) | 0.97<br>(0.90-1.04) | 1.01<br>(0.93-1.09) |
| Happy to be enrolled in current DSD model |  |  |  |  |  |  |  |
| No | 50 (5) | Ref | Ref | Ref | Ref | Ref | Ref |
|  |  | RD | ARD* | RD | ARD* | RD | ARD* |
| Yes | 999 (95) | 1.09<br>(1.01-1.16) | 1.05<br>(0.98-1.13) | 1.21<br>(1.14-1.28) | 1.06<br>(1.01-1.13) | 1.14<br>(0.99-1.32) | 1.12<br>(0.97-1.28) |
| **Facility location** |  |  |  |  |  |  |  |
| Rural | 494 (47) | Ref | Ref | Ref | Ref | Ref | Ref |
| Urban | 555 (53) | 1.03<br>(0.99-1.07) | 1.01<br>(0.97-1.06) | 0.97<br>(0.92-1.02) | 0.94<br>(0.90-0.99) | 1.01<br>(0.95-1.06) | 1.03<br>(0.97-1.10) |
| **Facility size** |  |  |  |  |  |  |  |
| 1000-2000 ART clients | 158 (15) | Ref | Ref | Ref | Ref | Ref | Ref |
| 2000-4000 ART clients | 424 (40) | 1.00<br>(0.94-1.06) | 1.04<br>(0.98-1.11) | 0.80<br>(0.74-0.88) | 0.91<br>(0.84-0.98) | 0.94<br>(0.87-1.03) | 1.00<br>(0.92-1.08) |
| 4000-6000 ART clients | 249 (24) | 1.05<br>(0.98-1.13) | 1.08<br>(1.00-1.16) | 0.88<br>(0.80-0.97) | 0.98<br>(0.89-1.06) | 1.00<br>(0.92-1.10) | 1.02<br>(0.94-1.12) |
| >6000 ART clients | 218 (21) | 1.03<br>(0.96-1.11) | 1.12<br>(1.04-1.22) | 0.70<br>(0.65-0.76) | 0.95<br>(0.87-1.03) | 0.90<br>(0.81-0.98) | 0.95<br>(0.85-1.06) |
| **DSD model** |  |  |  |  |  |  |  |
| Facility-based individual | 576 (550) | Ref | Ref | Ref | Ref | Ref | Ref |
| Facility-based group | 190 (18) | 0.98<br>(0.93-1.02) | 0.98<br>(0.93-1.04) | 0.98<br>(0.93-1.03) | 1.02<br>(0.96-1.08) | 0.88<br>(0.81-0.96) | 0.86<br>(0.79-0.95) |
| Community-based group or individual | 283 (27) | 1.16<br>(1.09-1.23) | 1.15<br>(1.08-1.22) | 1.42<br>(1.34-1.52) | 1.21<br>(1.12-1.30) | 1.23<br>(1.16-1.30) | 1.22<br>(1.14-1.31) |

\*Adjusted by DSD model, facility size, clinic locality, duration on ART, country, age, and sex.

Compared to clients in facility-based individual care models, clients enrolled in community-based models were somewhat more likely to have been offered a choice (1.21, [1.12-1.30]) and more likely to have asked for DSD model enrollment (1.15, [1.08-1.22]). Finally, those reporting that they were happy with their current model of care were likely to have been offered a choice (1.06, [1.01-1.13])

### Health provider study population

Characteristics of the 404 providers enrolled in the study are presented in Table 4. Consistent with the overall healthcare provider workforce, a majority were female. Most had been in their current positions for five years or more. Nurses at any level were most commonly enrolled in the study, particularly in South Africa, where they provide a large share of HIV care. In Malawi and Zambia, roughly half of the providers enrolled in the study were employees of partner organizations, rather than the Ministry of Health. In South Africa, the provincial departments of health employ most public-sector healthcare workers, including those enrolled in the study.

**Table 4.** Characteristics of health providers study population (n=404)

| Characteristics | Malawi | South Africa | Zambia |
| --- | --- | --- | --- |
| N (row percentage) | 110 (27) | 175 (43) | 119 (29) |
| Age (median, IQR) | 35 (31- 41) | 37 (31-50) | 34 (28-43) |
| Sex (female) | 68 (62) | 151 (86) | 73 (61) |
| Years in current role (median, IQR) | 5 (4-11) | 10 (5-15) | 6 (3-9) |
| Cadre |  |  |  |
| Healthcare Professionals | 85 (77) | 137 (78) | 86 (72) |
| Lay health workers | 11 (10) | 23 (13) | 21 (18) |
| Administrative, support and other staff | 14 (13) | 15 (9) | 12 (10) |
| Employer |  |  |  |
| Ministry/Department of Health | 65 (59) | 159 (91) | 58 (49) |
| Partner organization | 45 (41) | 16 (9) | 61 (51) |

### Health provider self-report of offering choice to ART clients

The majority of providers in all three countries stated that they do offer clients the option to choose between remaining in conventional care or enrolling in a differentiated model (Figure 2). Approximately 35% of providers in Zambia reported not offering this choice routinely, however, and a smaller proportion of providers said that they do not offer this choice occasionally, implying that they use personal judgment as to whether to make the offer to any individual client. Most providers also stated that they give clients an option among available DSD models, with this practice being most common in South Africa.

**Figure 2.**
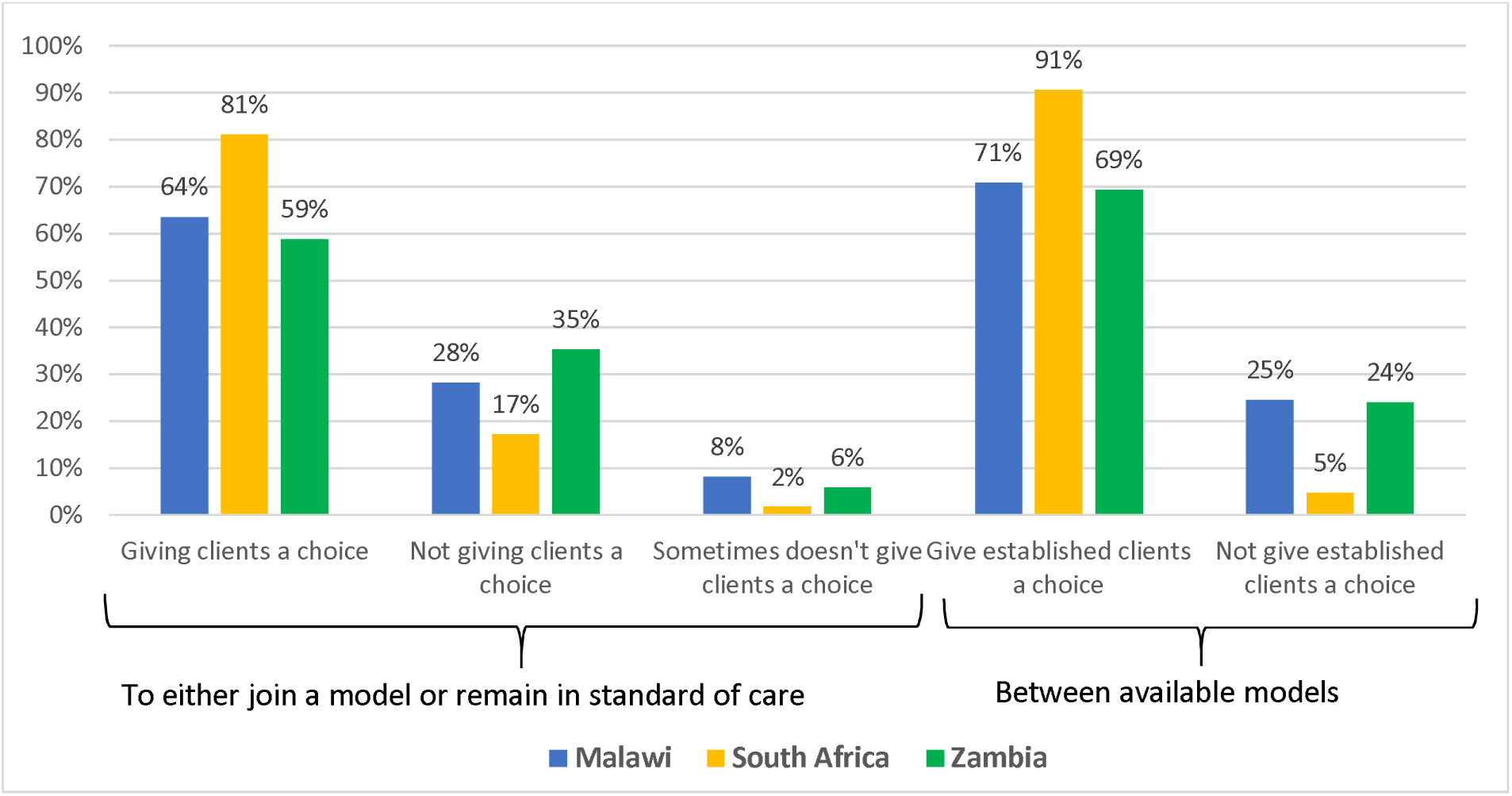
Health provider self-report of offer of choice in DSD model participation to ART clients, by country (n=404).

The majority (70%) of providers reported providing information about DSD models to all clients, regardless of the individual client’s DSD eligibility status (Figure 3). In all three countries, most of the information provided to patients about DSD models focused on eligibility criteria for enrolment, the frequency of visits, and the potential benefits of DSD models (Table 5).

**Figure 3.**
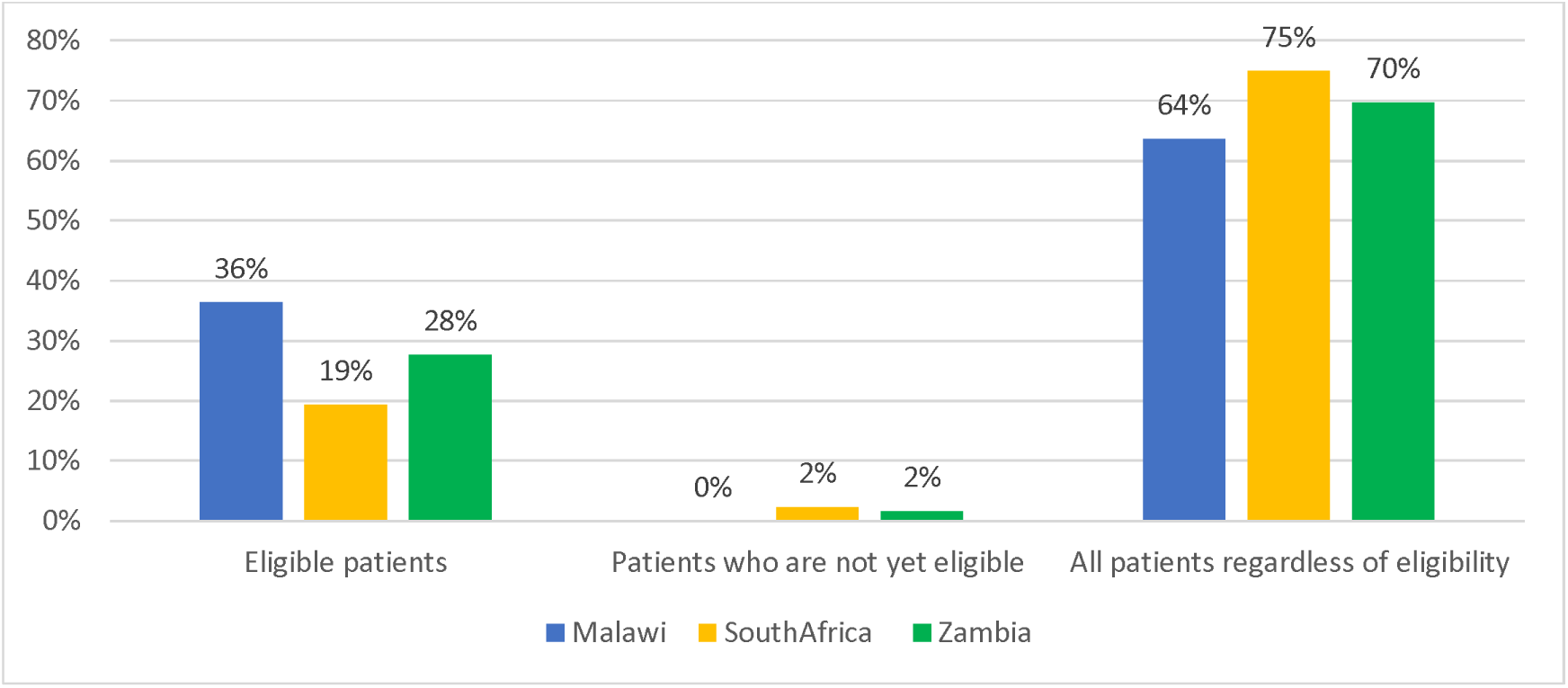
Clients to whom information about DSD models is conveyed, provider self-report (n=394).

**Table 5.** Information provided to patients on available DSD models (n=404)

| Information provided | Malawi | South Africa | Zambia |
| --- | --- | --- | --- |
| N (row %) | 110 (27) | 175 (43) | 119 (30) |
| Eligibility criteria | 102 (93) | 153 (87) | 113 (95) |
| Location of medication collection | 74 (67) | 144 (82) | 71 (60) |
| Type of provider seen | 64 (58) | 101 (58) | 49 (41) |
| Frequency of visits | 99 (90) | 141 (81) | 94 (79) |
| Potential benefits of the models | 91 (83) | 137 (78) | 99 (83) |
| Potential drawbacks of the models | 47 (43) | 58 (33) | 49 (41) |

Providers in Malawi, South Africa, and Zambia reported that clients are informed about DSD models through multiple mechanisms (Table 6). Health education talks occurring in the common areas of the clinic throughout the day were the most widely described mechanism to convey information about DSD models, benefits, and eligibility criteria. Providers frequently noted that these talks are reinforced during one-on-one consultations or counseling sessions between providers and clients which provides clients the opportunity to ask further questions. Additionally, a few providers specifically discussed sharing information through brochures given to clients or posters in common areas of the clinic. Emerging themes were not substantively different between countries.

**Table 6.** Information about DSD models offered at the facility, as reported by providers (n=282)*

| Key Themes | Illustrative Quotes |
| --- | --- |
| Health education talks which occur in the clinic common areas at least weekly | • “Weekly health talks where we explain everything about these models so that they should fully understand how they work.” - Malawi, female provider, age 32<br>• “We do health talks in the morning with all patients on ART.” - Malawi, male provider, age 25-34<br>• “Every day early in the morning [we] give them [patients] education about DSD models and the qualifications criteria to meet by each patient.” - South Africa, female provider, age 25-34<br>• “We give information through health education nearly every day to patients as they wait to be attended to.” - Zambia, female provider, age 35-49 |
| During routine consultations with providers | • “Providers give detailed information concerning DSD models to clients during consultation and health talks.” - Malawi, male provider, age 35-49<br>• “We do continuous sensitization from initiation and at each clinical visit.” - Zambia, female provider, age 25-34<br>• “Health education and health promoter talks to patients every morning and when they come to our consultation room, they would then ask more questions regarding the DSD models if they are interested.” - South Africa, female provider, age 50-64 |
| During adherence counseling sessions | • “Morning health talk, at the time of consultation and during counselling.” - Malawi, female provider, age 25-34<br>• “It is discussed during adherence counselling sessions and morning health talk before clinical sessions begin.” - Zambia, female provider, age 25-34<br>• “By telling them at different points. It's usually a part of adherence counselling especially on the benefit side.” - Zambia, male provider, age 35-49 |
| Through brochures and posters in clinic common areas | • “Health talks and printed information on the notice board at the clinic.” - Malawi, female provider, age 25-34<br>• “Through health education and pamphlets that are distributed here at the clinic.” - South Africa, female provider, age 35-49<br>• “Health education and educational material and wall posters. Also by one on one during consultation.” - South Africa, female provider, age 35-49 |

\*n = 84 for Malawi, n = 127 for South Africa, n = 71 for Zambia

When qualitatively explaining criteria for guiding clients on DSD model choices, providers in all three countries who responded (n=113, of whom 78% were from Malawi) discussed the official eligibility criteria for enrollment in DSD to first determine whether clients were eligible. Providers described offering choice to patients who were stable on treatment for at least six months and reported that the client’s home location was taken into account. For example, one provider in Zambia discussed that a group of patients who live close to each other might be enrolled in community adherence groups, while patients who live very far from the facility are recommended home delivery with 6MMD. Providers also discussed other criteria they might consider, such as age (teenagers often have separate DSD models), gender (if there are male-only models), comorbidities (which might affect the frequency or timing of clinic visits), and patients’ availability due to work or other obligations (to determine if it coincides with model requirements). As a crosscutting theme, providers discussed what they considered to be the right fit for the clients both within their ART treatment path and within their larger lives. Patient convenience and ability to benefit were key considerations (Table 7).

**Table 7.** Providers’ report of criteria they use for guiding patients on DSD model choice (n=110)*

| Key themes | Illustrative quotes |
| --- | --- |
| Viral load stability reflecting adherence to ART and to appointments; time on ART | • "Stable patients, with more than 6 months on ART being guided by MOH guidelines and criteria." - Malawi, male provider, age 25-34<br>• "Stable patients with suppressed Viral load, adherence, HIV stage. However, those with high Viral load and advanced HIV diseases can be enrolled in HVL and advanced disease respectively." - Malawi, male provider, age 25-34<br>• "We check how long has the client been on ART, age, health status and also viral load test." - Malawi, female provider, age 35-49<br>• "Patients stability- must have a suppressed viral load, good history of drug adherence and must have stable/ managed co-morbidity." - Zambia, male provider, age 25-34 |
| Patient's location | • "Client health condition, people who sometimes live far from facility are offered community outreach and special needs as for teen club." – Malawi, male provider, age 35-49<br>• "The area where the patient is staying, blood results, and adherence." - South Africa, female provider, age 35-49<br>• "We look at stability of the patient and where they come from. Those that stay near each other are usually put in CAGs [Community Adherence Groups] and those that come from very far are usually put on home delivery and 6MMD." - Zambia, female provider, age 50-64<br>• "We look at history of clients and where they come from. We prioritize those that come from far place e.g. [individuals living in village name are best] for |
|  | home delivery because they have challenges with transport.” - Zambia, male provider, age 25-34<br>• “We look at their VL and frequency of visits. We also consider location of patients” - Zambia, male provider, age 35-49 |
| Other criteria such as comorbidities, age, gender, availability, history | • “For adults, it depends on their adherence and viral load results. For young ones, it depends on their age and their understanding of the disease.” - Malawi, male provider, age 25-34<br>• “Adherence to ART, available of patient on specific days which DSD models are conducted.” - Malawi, male provider, age 25-34<br>• “We explore how long the client has been on treatment, client's adherence to treatment, viral load suppression, sex, age and then we decide a model for the client.” - Malawi, male provider, age 25-34<br>• “Age, patient’s choice, viral load and other chronic diseases.” – South Africa, female provider, age 35-49<br>• “We check for viral load and also the age of the client then that will help us determine the mode suitable for a client.” – Zambia, female provider, age 25-34 |

\*n=88 for Malawi, n=4 for South Africa, n=18 for Zambia

Providers were asked to generate ideas to improve DSD and client choice; emerging themes and illustrative quotes are presented in Table 8. Providers in all countries widely recommended providing clients with more information on the benefits and guidelines for DSD models to ensure they can make an informed choice. Providers discussed relaying this information to clients through multiple sources, including during counseling sessions, during general health education talks, through posters, on TV screens in the waiting areas, and in leaflets. A few providers noted the importance of this information being relayed in the primary language the patient spoke (e.g. Bemba in Zambia). Additionally, numerous providers described the importance of making this information available throughout the early treatment period, not only after the client becomes stable and eligible for enrollment in the DSD models. They speculated that early enrollment in DSD models could be motivating for good adherence if clients are informed of the benefits and eligibility requirements early in their treatment journey. Providers also recommended providing additional adherence support to clients so they can more quickly be eligible for DSD, including increased adherence counseling, outreach (SMS (text message), phone calls, or home visits if needed) after missed appointments, and interactions with peer educators.

**Table 8.** Key provider suggestions for facilitating patient choice of DSD models (n=370)*

| Suggestions | Illustrative quotes |
| --- | --- |
| Provide patients with more information on benefits and eligibility criteria for DSD models | • “During counselling, education and health talks, providers must explain in detail the importance and disadvantages of each model and then let the client make a choice of the model.” – Malawi, male provider, age 35-49<br>• “The screen in the waiting area should show information on DSD's and their eligibility criteria thereof. There should also be leaflets on the same for those who can read.” - Malawi, female provider, age 35-49<br>• “One of the better ways could be having peers to be explaining the benefits of having to be in a DSD model to patients as a way of encouraging them to take their medication serious and making sure they stay stable.” - Zambia, male provider, age 35-49<br>• “We can explain in detail through health talks the benefits of being in a respective DSD model and also explain the eligibility criteria and potential drawbacks so that clients can make informed decisions.” - Zambia, female provider, age 50-64 |
| Provide more adherence counselling, education, and support to patients | • “Adherence counselling support to clients [so patients do] not miss appointments for no good reasons.” – Malawi, male provider, age 25-34<br>• “Explain what DSDs are to clients for motivation. High viral load patients can be informed of 6MMD model which can encourage them on good adherence for suppressed viral load.” – Malawi, male provider, age 35-49<br>• “Patients must be encouraged to adhere to treatment by supporting them vigorously in terms of follow-up phone calls or by home visits or creating community group that will enable people to be supported even at community levels.” - South Africa, female provider, age 35-49<br>• “Maybe more sensitization in the community is needed to help the clients who are on ART to understand the advantage of good adherence. Only when we have more improved adherence can we be able to give patient choice.” – Zambia, female provider, age 35-49 |
| Give patients choice or decision-making power/ offer patients advice or guidance | • “There should be an understanding between the patient and the provider so that the patient is able to make their own choice.” – Malawi, male provider, age 25-34<br>• “To give clients enough information to make the right choice. You can also advise them based on the choice they make. [Even] if you see they do not realize the potential drawbacks of their choice, the final choice lies with them.” - South Africa, female provider, age 35-49<br>• “We don’t need to impose [the choice on them]. We need to counsel them and explain the models to them. Then they can choose which model best suits them, we can analyze the model to ensure that the model chosen by the patient is suitable for them.” – Zambia, female provider, age 50-64<br>• “By asking the patients and giving them much authority to decide how they want to receive the service.” – Zambia, female provider, age 32 |
| Create more DSD models or more options | • “New DSD models should be introduced as to include clients that are free on Sundays.” – Malawi, male provider, age 35-49<br>• “If we can have lots of pickup points in the community for patients to choose from.” - South Africa, female provider, age 35-49<br>• “Extend pickup points out of provinces so that wherever patients go they can produce their identity number and get treatment.” – South Africa, female provider, age 25-34 |
|  | • “Introduce more DSD models like Community Adherence Groups, fast tracking those just coming for medical pickup.” – Zambia, female provider, age 35-49 |
| Build capacity among providers | • “More trainings [to providers] should be provided to improve service delivery.” – Malawi, female provider, age 25-34<br>• “At this facility there are only two people who are working with DSD patients, so I think they should train more staff.” – South Africa, female provider, age 25-34<br>• “Introduction of staff training on DSD models. When the provider understands the different DSD models being offered, they are able to explain to the patients on the different DSD models that are being offered and the benefits of each models thus facilitating their patient choices.” – Zambia, male provider, age 25-34 |

\*n=108 for Malawi, n=150 for South Africa, n=112 for Zambia

During the conversation in which the client needs to choose their DSD mode, providers discussed the importance of offering complete information to the patients, including the benefits and drawbacks of each model. Many providers described either advising the patient on which model might be best suited for their needs or helping them to determine this, such as by identifying which external pickup point is closest to their home. Many also stated that it is just the client’s place to choose, and it wasn’t clear from their responses whether those providers offered additional decision-making support.

Some providers suggested various forms of expanded DSD enrolment, with more patients on DSD models, more DSD models available, or more medication pickup location options.

A small number of providers suggested additional and continuous capacity building and education for the providers so they can better explain the model options to clients and assist in this decision-making support role.

## Discussion

This mixed-methods study explored patient and provider perspectives on one element of client-centeredness of HIV treatment, the opportunity to choose a model of care. As of early 2023, 4% of HIV treatment clients enrolled in differentiated service delivery models in Malawi, 17% in Zambia, and 47% in South Africa reported being offered a choice of model prior to enrolment. Only a few in any of the focus countries said that they had asked to be enrolled in any DSD model, rather than remaining in conventional care, and over a third said that they would have preferred to be in a different model. On the other hand, roughly two-thirds reported consenting to DSD enrollment, and nearly all—between 91 and 99%—said that they were happy to be enrolled in their current model. Quantitative and qualitative data from providers (Figure 2, Table 8) suggest that they strongly endorse the principles of client-centered care and even recommend that DSD models and choice be introduced earlier in a patient’s treatment journey as a strategy to improve adherence. In contrast to clients’ perceptions of their experience, most providers in all three countries reported they did usually offer clients the choice either to enroll in a DSD model or to remain in conventional care to clients.

Our survey suggests a disconnect between what providers believe they are offering, in terms of both the opportunity for choice and the greater value of their own judgement, and what clients are experiencing with regard to choice.^13–15^ Qualitatively, most participants said that they were given only one option, to enroll in a specific model or remain in conventional care. Client-centered care is a fundamental tenant, if not the raison d’etre, of the differentiated service delivery approach. ^5^ Choice is considered a core element of patient empowerment and, thus, of client-centered care, which in turn has been shown to improve patient adherence and outcomes ^16–19^, including for HIV. ^17,20^ To the extent that the importance of choice pertains to our study setting, DSD can only achieve its goals if patients have the opportunity to make their own decisions, where options are available. In our analysis (Table 3), we found that having been offered a choice was the factor most consistently associated with self-reported happiness to be in the client’s DSD model.

At the same time, the findings reported here suggest that being offered choice is not essential to overall, self-reported patient satisfaction with care received. Patients may find that the services they receive meet or exceed their expectations regardless of having had a choice, or they may be unaware of alternative options, leading to satisfaction by default rather than by preference. ^14^ It is possible that clients simply prefer any DSD model to conventional care and/or that they had no expectation of being empowered to participate in decisions. Alternatively, some clients may prefer an “opt-out” approach to DSD models, in which they are enrolled in a model of the provider’s choice unless they actively express a different preference, in order to simplify clients’ lives, mitigate the burden or anxiety associated with having to make a choice, or reassure the clients of the expertise of the provider.^15^

In Malawi and Zambia, many facility-based DSD models limit enrollment based on individual characteristics such as gender (e.g. male adherence clinic), age (teen/youth clubs), current life stage such as being pregnant/post-partum (mother-infant pairs or family model clinic), or clinical condition (not being established on ART for a high viral load clinic). These requirements constrain the choices available to some clients, such that for some individuals at some study sites, only one DSD option would have been both available and suitable. These limitations may have led healthcare providers to recommend only one specific model and may explain why study participants reported having less choice in these countries, a speculation supported by our qualitative results. Where more than one alternative was available, though, and clients were still assigned to a model based on their characteristics without being given a genuine opportunity to choose, then enrollment may not have truly reflected a personal preference. ^21–24^ Providers in Malawi and Zambia may thus have an opportunity to improve outcomes by offering sufficient information about all models a client is eligible for (including remaining in conventional care) rather than solely the specific model that matches that individual’s characteristics. Actively involving the client in the decision-making process may also improve treatment outcomes ^25,26^, for example, by imbuing a sense of responsibility for self-management of care. ^27^

An important finding of this study is that providers’ explanations of how they determine which DSD model(s) to recommend diverge from guidelines to some extent and clearly involve individual judgment (Table 6). The patient’s location, for example, was frequently cited as a reason for recommending one model over another, implying that distant patients may be offered community-based models like home delivery more frequently than those who live nearby. Location is not a criterion mentioned in DSD guidelines. More important, both distant and nearby patients may have other considerations that take priority for them as individuals, such as fear of disclosure to neighbors due to home delivery or a community adherence group. The provision of full information about all available models, without undue pressure to select the model recommended by the provider, may help offset this concern. Our study had several limitations. With regard to patient results, response and recall biases, where participants may inaccurately report or remember details about the choices offered, are likely, as many participants were describing experiences that occurred several years in the past. Respondents may have had different interpretations of what constitutes a meaningful choice, and the influence of healthcare providers in presenting options could skew perceptions. Our study only included patients enrolled in DSD models; those who remained in conventional care at the time of the survey were excluded, even though many of them may have chosen the conventional care model. As a result, the study may not accurately reflect how choices are offered or perceived by patients who prefer or are recommended to remain in conventional care.

On the provider side, respondents may have overstated how often or effectively they offer choices to their ART clients. Recall bias could affect the accuracy of their recollections of client interactions, and variability in how they interpret the concept of offering a choice could lead to inconsistent responses. Social desirability bias may also cause providers to report practices that align with expected norms rather than their actual behaviors. Finally, the cross-sectional nature of the study also limits its ability to capture changes in provider practice or client experience over time and could also affect the reliability and generalizability of the findings.

## Conclusion

Differentiated service delivery for HIV treatment has been widely implemented throughout sub-Saharan Africa and is generally considered a success in terms of clinical outcomes and patient satisfaction. One of its main goals—in fact, its primary goal--is to increase the client-centeredness of HIV treatment, which in turn entails offering clients the opportunity to choose among existing options for service delivery models. In this study we found that relatively few ART clients in Malawi, South Africa, and Zambia in 2023 reported they were offered such a choice, despite most providers having reported almost always offering DSD information and choice. Further examination of what clients and providers consider to constitute “choice” and improving healthcare provider communication with patients could improve client-centeredness. Other potentially important questions to answer in future research include the value of choice in improving both clinical and non-clinical outcomes, facilities’ capacity to offer choice and whether there is a quantity/quality tradeoff in having multiple options, whether choices evolve over clients’ lifetimes, and how to make the choice process dynamic over time.

## List of abbreviations

DSD: Differentiated service delivery
ART: antiretroviral therapy
HIV: human immunodeficiency virus
DMOC: Differentiated Models of Care
RPC: Repeat Prescription Collection Strategies
COVID-19: coronavirus disease 2019
MMD: multi-month dispensing
IQR: interquartile ranges
RD: risk differences
ARD: adjusted risk differences
CI: confidence intervals
CHW: Community Health Workers
SMS: short message service

## Declarations

### Ethics approval and consent to participate

Ethical approval to conduct this study was granted by University of Witwatersrand (Medical) Human Research Ethics Committee in South Africa (Protocol M210241), the National Health Science Research Committee (NHSRC) in Malawi (protocol 21/03/2672), ERES Converge Institutional Review Board in Zambia (Protocol 2021-Mar-012), and by the Boston University Medical Campus Institutional Review Board in the United States (Protocol H-41402). Data collectors were trained in research ethics, the overarching study, and the specific survey instrument. Written informed consent was obtained from each participant before the survey commenced.

### Consent for publication

Not applicable.

### Data availability

All data used in this study were collected by the study team following written informed consent. Data will be made available within one year after the closure of the study by the supervising ethics committees. At that time, data will be posted in a public data repository.

### Authors’ contributions

SR, AH, IM and SP conceptualized the study. IM, SR, AH, SP, TT, AK and WP contributed to instrument design. IM, VN, JLK, NS, OM and AM analyzed the data. IM, NS, and SR drafted the manuscript. PH, HS, PLM and RN provided feedback on the manuscript. All authors reviewed and approved the final manuscript.

## Acknowledgements

We would like to express our gratitude to the staff at the study sites for their participation in the research. Additionally, we extend our thanks to the Malawi Ministry of Health, the South Africa National Department of Health, and the Zambia Ministry of Health for approving our research. We are especially thankful for the support provided by the Differentiated Service Delivery Technical Working Group in each country. We would also like to acknowledge the other members of the AMBIT SENTINEL study team for their contributions to the project at large: Frehiwot Birhanu, Nyasha Mutanda, Linda Sande, Nkgomeleng Lekodeba, Nkosinathi Ncgobo, Aniset Kamanga, and Taurai Makwalu.

## Supporting information

**Supplementary Table 1. Models of care commonly offered at study sites**

**Supplementary File 1. SENTINEL patient survey instrument**

**Supplementary File 2. SENTINEL provider survey instrument**

