## Supplementary Table 1 for "Are HIV treatment clients offered a choice of differentiated service delivery models? Evidence from Malawi, South Africa, and Zambia"

### Supplementary Table 1. Models of care commonly offered at study sites

| Model | Description |
|---|---|
| **Malawi** | |
| Conventional care | Clients receive a 3-month supply of medications at each full clinic visit. |
| Six-month medication dispensing (6MMD) | Clients receive a 6-month supply of antiretroviral medications at each full clinic visit |
| Mother-infant pair | The post-partum visits for the mother are aligned to the infant visit schedule. The infant's schedule is based on the vaccination milestones. Thereafter the mother receives a 3-month supply of antiretroviral medications at each visit |
| Teen Club | A support group initiative for adolescents living with HIV who are aware of their HIV status, conducted on a monthly or bi-monthly basis, on a Saturday or Sunday, outside of normal clinic hours. |
| Family model clinic | A family-centered approach of providing integrated HIV care and treatment for entire families, including husbands, wives, and children who are HIV positive and on treatment. |
| Community ART distribution (CADs) | Provider-led in which provider–led ART teams deliver integrated HIV services at health posts in communities on scheduled days |
| Community ART Group (CAGs) | ART-client -led model for ART distribution, whereby groups of PLHIV rotate for clinic visits and drug refills at the clinic while dispensing drugs to their peers in the community and ensuring peer support. |
| Advanced HIV Disease clinic | The model aims to meet the needs of people living with advanced HIV disease, particularly those with low CD4 counts, high viral loads, or opportunistic infections. It focuses on individuals at higher risk of health complications and mortality due to late presentation or failure to adhere to treatment. |
| **South Africa** | |
| Conventional care | Clients receive a 1-2-month supply of medications at each full clinic visit. |
| Facility-based pickup points | Between full clinic visits, clients pick up medications (usually a 2-3-month supply) at specified pickup points in facilities. |
| External pickup points | Between full clinic visits, clients pick up medications (usually a 2-3-month supply) at specified pickup points in the community (e.g. commercial pharmacy) |
| Adherence clubs | Facility or community-based group model that is led by a health care worker (professional or lay) where clients pick up medications (usually a 2-3-month supply) |
| Home ART delivery | A model that delivers ART to patients' homes (e.g., by a community health worker or a bicycle courier) (usually a 2-3-month supply) |
| **Zambia** | |
| Conventional care | Clients receive a 3-month supply of medications at each full clinic visit. |
| Six-month medication dispensing (6MMD) | Clients receive a 6-month supply of antiretroviral medications at each full clinic visit. |
| Fast track | Clients go directly to the facility's dedicated Fast Track room to receive a 3-6-month supply of medication. The model also includes an appointment system, where clients and providers agree on a specific day and time for the next visit. |
| Teen treatment clubs | A youth-centered care model where adolescents typically meet every 1-3 months at health facilities, youth-friendly spaces, or community centers to receive their pre-packed ART, undergo brief health check-ups, and participate in club activities. |
| Health post | ART is pre-packed for each client at the healthcare facility and then delivered to the health post, where a 3-6-month supply is dispensed. Clients only need to visit the healthcare facility once every 6 months for their clinical review and laboratory monitoring. |
| Community Post | The use of private retail pharmacies as designated points for ART distribution (usually a 3-6-month supply). This approach aims to make ART more accessible and convenient, particularly for stable patients who do not need frequent clinical monitoring. |
| Community ART Group (CAGs) | Clients form groups of 2-6 members and receive their ART refills during monthly or quarterly community meetings. The group is self-managed, with members taking turns collecting medication from the health facility. |
| Community ART access points (CAAPs) | A lay worker collects 3-month supply of medication for 8 clients and distributes it at a designated CAAP. |
